# Development and Validation of a Diagnostic Prediction Rule for Osteopenia

**DOI:** 10.1101/2024.05.23.24307788

**Authors:** Thammabhorn Janwittayanuchit, Naritsaret Kaewboonlert, Pornthep Tangkanjanavelukul, Kitirat Phattaramarut, Pattama Thongdee

**Author notes:** Corresponding author: School of Surgery, Institute of Medicine, Suranaree University of Technology, Nakhon Ratchasima 30000, Thailand.

## Abstract

**Objectives:** To triage patients with a high likelihood of osteopenia before referring them for a standard bone mass density test for diagnosis.

**Introduction:** Osteopenia defined by low bone mineral density, is a precursor for osteoporosis and is primarily associated with aging-linked natural bone loss in adulthood. The model and findings can be used to adopt an inclusive screening and swift treatment model that can work in most settings where resources are limited.

**Methods:** We developed a diagnostic prediction rule based on clinical characteristics. A retrospective cohort of 798 patients who were going to be diagnosed with osteopenia or osteoporosis, within January-September 2022. The multivariable logistic regression to assess potential predictors. The logistic coefficients were transformed as a risk-based scoring system. The internally validation was performed using a bootstrapping procedure.

**Results:** The model initially included seven predictors: sex, age, height, weight, body mass index, diabetes mellitus, and estimated glomerular filtration rate. However, after using backward elimination for model reduction, only three predictors—sex, age, and weight—were retained in the final model. The discrimination performance was assessed with the area under the receiver operating characteristic curve (AuROC); it was 0.779 (95%CI 0.74-0.82), and the calibration plot showed good calibration. For internal validation, bootstrap resampling was utilized, yielding an AuROC of 0.768 (95% CI 0.73-0.81), indicating robust performance of the model.

**Conclusions:** This study developed and internally validated the Osteopenia Simple Scoring System. This clinical risk score could be one of the important tools for diagnosing osteopenia and allocating resources in resource-limited settings.

## Introduction

Osteopenia is a decrease in bone mineral density (BMD) below normal values for their ages, is the initial stage of bone loss, which may progress to a more severe condition i.e. osteoporosis. However, that doesn’t always lead to osteoporosis depending on many factors. The primary cause of osteopenia is the natural bone loss that occurs gradually during adulthood. Secondary causes supposed to accelerate bone loss include lifestyle factors[1] such as smoking, certain underlying diseases, steroid usage, early menopausal woman, rheumatoid arthritis, and some medications as well. Osteopenia is often a precursor to osteoporosis, which are now diagnosed by measuring bone mineral density using dual energy X-ray absorptiometry bone scans.[2] The osteopenia, as defined by the World Health Organization (WHO) is a t-score between −1 to −2.5, while values less than −2.5 are diagnostic for osteoporosis.[3, 4] Osteopenia is not considered a disease while osteoporosis is. In the other hand, osteopenia is considered a marker for risk of fractures.[5]

The potential predictors of bone mass density in patients with fractures treated in hospitals were found that factors such as age, sex, smoking, history of adult wrist fractures, spinal deformities[6, 7], history of adult hip fractures, and osteoarthritis of the spine[8] significantly differed statistically between groups with normal bone mass density and those with bone mass density below −1 standard deviation. Low body mass index, low vitamin D level[9] and diabetes[10, 11], chronic kidney disease[12] are also associated with osteoporosis.

There has been increasing attention to the clinical predictive models of diagnostic screening models for the prediction of fracture risk in patients diagnosed with osteoporosis.[13–16] Clinical predictive models are commonly used in clinics for the purpose of disease diagnosis, outcome prediction, and evaluation of the clinical response.[17, 18] We used multivariable logistic regression to develop predictive models for possible use in the facilitation of early treatment and screening for osteopenia.

In countries with limited resources, access to a test for bone mass density would be far-fetched. This research aims to help triage patients at high risk of having osteopenia before they are referred for a standard BMD test for diagnosis. This work could be extended to programs aimed to osteopenia or osteoporosis screening in the community.

## Methods

### Study design and setting

This diagnostic prediction research, utilizing a retrospective cohort design, was conducted at Suranaree University of Technology Hospital in Nakhon Ratchasima, located in the lower northeastern region of Thailand. Our university hospital conducts more than 2,000 bone mass density tests annually.

This study retrospectively collected demographic and laboratory data from the electronic medical records of Suranaree University Technology Hospital. The data included age, weight, height, body mass index, smoking status, early menopause, rheumatoid arthritis, diabetes mellitus, chronic kidney disease, serum creatinine levels, and estimated glomerular filtration rates. Patient data were accessed for research purposes between April 4th and 12th 2024. Patients who visited the hospital from January to September 2022 and were intended to be diagnosed with osteopenia or osteoporosis were assessed for eligibility. Patients with a previous diagnosis of osteopenia or osteoporosis were excluded from the study.

### Confirmation of cases

All patients included in this study underwent a BMD test on a single machine at the hospital’s checkup center. In this study, osteopenia is defined as a T-score of less than −1, and osteoporosis is defined as a T-score of less than −2.5.

### Statistical analysis and sample size calculations

Continuous variables were assessed for normality and presented as means and standard deviations if normally distributed, or medians and interquartile ranges if not. Mean differences of the variables between the two groups were compared by using an independent t-test, or rank-sum test based on the distribution of the data. Categorical data were expressed as frequencies and percentages of the total in each group and compared between groups using either the exact probability test or chi-square test where applicable. We assessed diagnostic performance and potential prediction by univariable logistic regression, using crude odds ratios (OR) and their corresponding area under the receiver operating characteristic curves (AuROC). Statistically significant two-sided p-values less than 0.05 were considered in applicable cases. All analyses were done using Stata statistical software version 17. In the development of clinical prediction rules based on methods described by Riley et al.[19], it was estimated that the minimum sample size for a multivariable prediction model with a binary outcome was required. This was estimated from a model c-statistic of 0.8, six candidate predictors, and an assumed prevalence of osteopenia from a preliminary study, standing at 46%, to get the minimum sample size of 382 cases with 176 events.

### Model development and validation

All potential predictor variables necessary for diagnostic prediction of osteopenia in our routine practice were extracted from the hospital’s electronic medical records. These included age, sex, height, weight, smoking status, serum creatinine levels, diabetes mellitus (with or without insulin use), hypertension, early menopause, chronic kidney disease, steroid use, and rheumatoid arthritis.

We identified potential predictors based on prior knowledge; about the biological process, a review of the literature, and available prediction models. Subsequently, the exploratory analysis of significant predictors was done using a univariable logistic regression. We assessed the significance of the predictors through the diagnostic odds ratio and the corresponding p-value. Additionally, we assessed the area under the receiver operating characteristic curve (AuROC) for each univariable logistic model. Any predictor variable showing an odds ratio >1.00, significant p-value of <0.05, and higher AuROC than others was included in the model. Continuous potential variables were categorized into ordinal following the preceding model and review of literature. Therefore, in this respect, understanding the nature of the relationship between the dependent variable and the outcome to determines the cut-off point.

The model to be used for the study is derived from the multivariable logistic regression with a binary outcome. The factors that not contributing to the outcome were removed using the backward elimination method. A total of four predictors got pruned from the model: height, body mass index, diabetes mellitus, and estimated glomerular filtration rate. Diagnostic performance of the developed model was assessed using the reduced multivariable model by means of calibration and discrimination. Calibration was assessed by the Hosmer-Lemeshow goodness-of-fit test, and a plot was applied to show both the model-estimated disease probabilities and observed disease data. The discriminative ability of the model was graphically tested through a distributional plot. It was reported with the area under the curve of the receiver operating characteristic. The internal validation was conducted using the bootstrapping procedure of 1000 replications.

### Simplified score derivation

Each predictor in the multivariable model was assigned a specific score based on the logistic regression coefficients. The coefficient of each predictor was divided by the smallest coefficient and then rounded up to the nearest whole number. Utilizing a population-analogue approach, the positive predictive value (PPV) was calculated to demonstrate the predictive performance. Calibration and discrimination measurements were also conducted using the score-based multivariable logistic model.

### Ethical considerations

This research was conducted based on ethical standards of clinical research. According to the Helsinki Declaration and began its activity only after it received approval and permission from the Institutional Review Board of Suranaree University of Technology regarding the review of the research protocol. Retrospective data were extracted through data record forms. The patients were treated by the routine hospital staff and were not affected by any research protocols, the informed consent was waived. The study adhered to the reporting guidelines outlined in the Transparent Reporting of a multivariable prediction model for Individual Prognosis Or Diagnosis (TRIPOD) statement. The study protocol received approval from the Institutional Review Board of Suranaree University of Technology, with approval number COA No.32/2567.

## Results

### Participants

A total of 798 participants were evaluated for osteopenia at the Suranaree University of Technology check-up center from January to September 2022. After excluding 242 patients previously diagnosed with osteopenia or osteoporosis, the remaining 556 patients were divided into two groups: 230 in the osteopenia group and 326 in the non-osteopenia group. The prevalence of osteopenia and osteoporosis was 41.4% and 5.4%, respectively. Of these, 198 patients in the osteopenia group and 188 in the non-osteopenia group were female. The mean age for the osteopenia group was 65.07±10.34 years, compared to 59.03±9.14 years for the non-osteopenia group. The average weight was 57.40±9.58 kg for osteopenia cases and 57.40±12.71 kg for non-osteopenia cases. Average heights were 155.22±7.50 cm for the osteopenia group and 160.53±8.00 cm for the non-osteopenia group. The mean BMI was 24.68±14.44 kg/m^2 for osteopenia cases and 25.64±4.02 kg/m^2 for non-osteopenia cases (Table 1). There were significant differences in groups in terms of female gender, age, weight, height, BMI, and current underlying diseases, including diabetes mellitus, as well as in laboratory factors such as estimated glomerular filtration rate (eGFR) (p < 0.001). There were no significant differences in the serum creatinine, early menopause, rheumatoid arthritis, and smoking status.

**Table 1:**
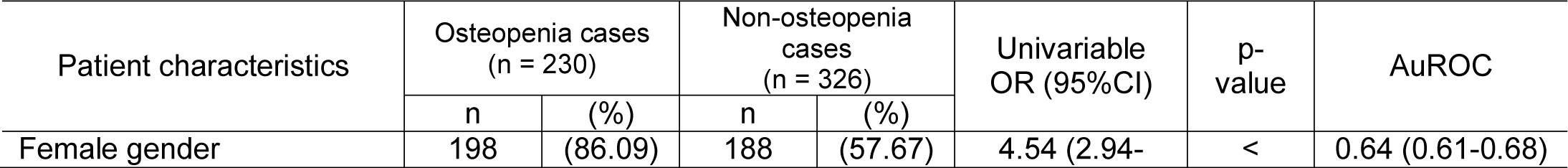
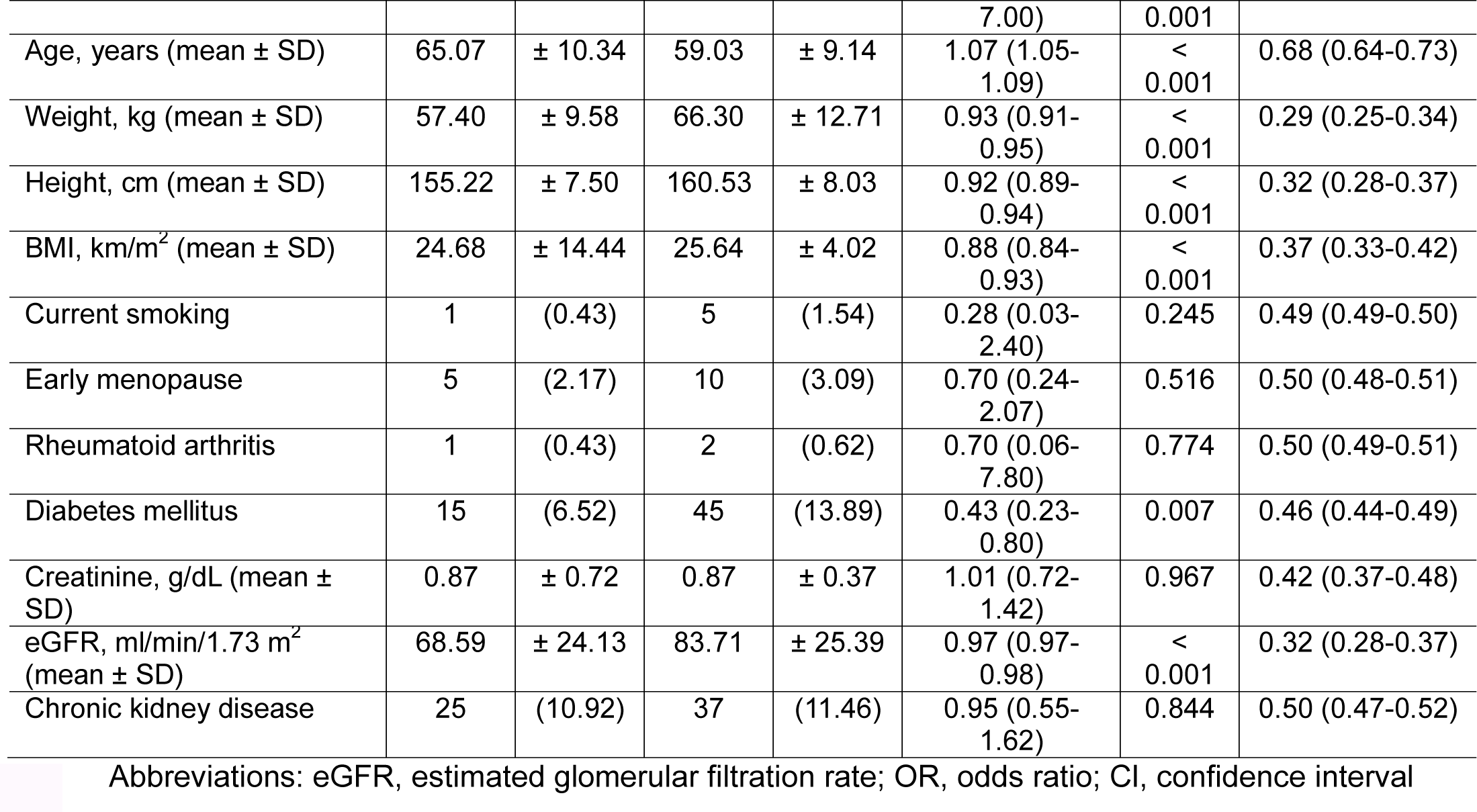
Baseline patient characteristics, underlying diseases, and laboratory investigations of the derivation cohort, along with a comparison of osteopenia cases and normal bone mineral density tests (n = 556).

### Model development and internal validation

The following continuous variables were converted to ordinal variables: eGFR, body mass index, weight, age, and height. The cut points were determined based on information gathered from the literature and prior models. The model was developed using multivariable logistic regression, which demonstrated the relevant characteristics included in the regression for predicting osteopenia were female gender, age, height, weight, BMI, diabetes mellitus, and eGFR. These factors remained significant in the multivariable logistic regression analysis and were integrated into the full risk prediction model (Table 2).

**Table 2:** Full model multivariable logistic regression analysis.

| Predictors | Full model |  |  |
| --- | --- | --- | --- |
|  | mOR | 95% CI | P-value |
| Female | 5.33 | 3.03 - 9.38 | 0.000 |
| Age, year |  |  |  |
| ≤ 59 | 1 |  |  |
| 60 – 69 | 3.59 | 2.25 - 5.74 | < 0.001 |
| ≥ 70 | 6.91 | 3.62 - 13.20 | < 0.001 |
| Height, cm |  |  |  |
| ≥ 155 | 1 |  |  |
| < 155 | 0.97 | 0.57 - 1.64 | 0.903 |
| Weight, kg |  |  |  |
| ≥ 70 | 1 |  |  |
| 60 – 69 | 1.15 | 0.61 - 2.17 | 0.656 |
| 50 – 59 | 1.66 | 0.72 - 3.82 | 0.233 |
| ≤ 49 | 1.72 | 0.54 - 5.54 | 0.360 |
| Body mass index, kg/m<sup>2</sup> |  |  |  |
| ≥ 25.0 | 1 |  |  |
| 23.0 – 24.9 | 1.19 | 0.64 - 2.22 | 0.577 |
| 18.5 – 22.9 | 1.71 | 0.79 - 3.69 | 0.169 |
| < 18.5 | 3.94 | 0.87 - 17.90 | 0.076 |
| Diabetes mellitus | 0.35 | 0.17 - 0.71 | 0.004 |
| Estimated GFR |  |  |  |
| ≥ 60 | 1 |  |  |
| 30 – 59 | 0.84 | 0.45 - 1.54 | 0.565 |
| 15 – 29 | 1.89 | 0.22 - 16.05 | 0.558 |
| ≤ 14 | 1 | Empty |  |
| Model intercept | .062 |  |  |

Abbreviation: mOR, multivariable odds ratio; BMI, body mass index; GFR, glomerular filtration rate

The reduced model employed a backward elimination strategy. Multivariable logistic regression indicated that female gender, age, and weight, which were included in the final model, were found to be statistically significant. The scoring ranged from 1 to a maximum of 5 points, accumulating to a total of 13 points, summing up to 133. Ages over 70 years were assigned the highest scores (Table 3).

**Table 3:**
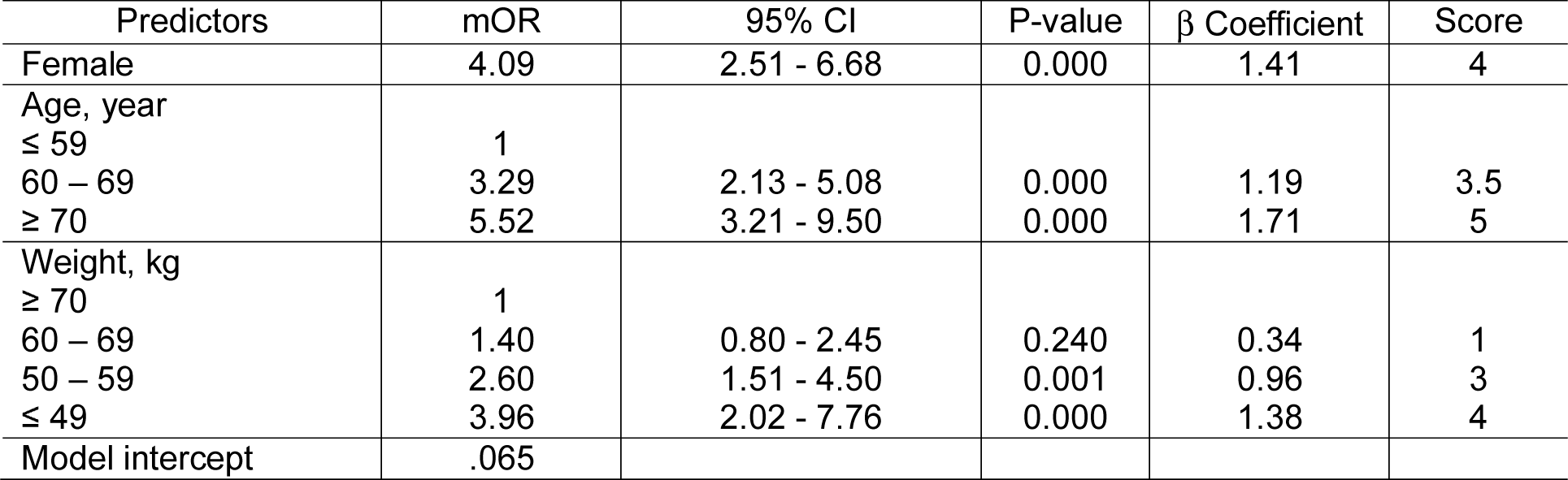
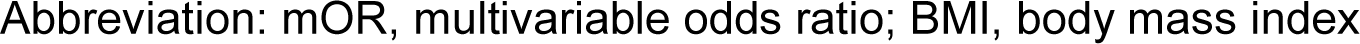
Reduced model with logit coefficients.

| Predictors | mOR | 95% CI | P-value | β Coefficient | Score |
| --- | --- | --- | --- | --- | --- |
| Female | 4.09 | 2.51 - 6.68 | 0.000 | 1.41 | 4 |
| Age, year |  |  |  |  |  |
| ≤ 59 | 1 |  |  |  |  |
| 60 – 69 | 3.29 | 2.13 - 5.08 | 0.000 | 1.19 | 3.5 |
| ≥ 70 | 5.52 | 3.21 - 9.50 | 0.000 | 1.71 | 5 |
| Weight, kg |  |  |  |  |  |
| ≥ 70 | 1 |  |  |  |  |
| 60 – 69 | 1.40 | 0.80 - 2.45 | 0.240 | 0.34 | 1 |
| 50 – 59 | 2.60 | 1.51 - 4.50 | 0.001 | 0.96 | 3 |
| ≤ 49 | 3.96 | 2.02 - 7.76 | 0.000 | 1.38 | 4 |
| Model intercept | .065 |  |  |  |  |

Abbreviation: mOR, multivariable odds ratio; BMI, body mass index

The calibration plot of the estimated risk of osteopenia compared to the actual risk showed acceptable calibration, with observed probabilities closely matching the expected probabilities and exhibiting minimum variation from the ideal (Figure 2). The Hosmer-Lemeshow goodness-of-fit statistic yielded a non-significant result for the outcome (p = 0.614), suggesting that the statistical fitness of the model was satisfactory, given that a p-value larger than 0.1 was regarded indicative of a good fit. Area under the receiver operating characteristic curve (AuROC) was 0.7792 (95% CI 0.74-0.82), showing a good performance in model discrimation (Figure 3).

**Figure 1:**
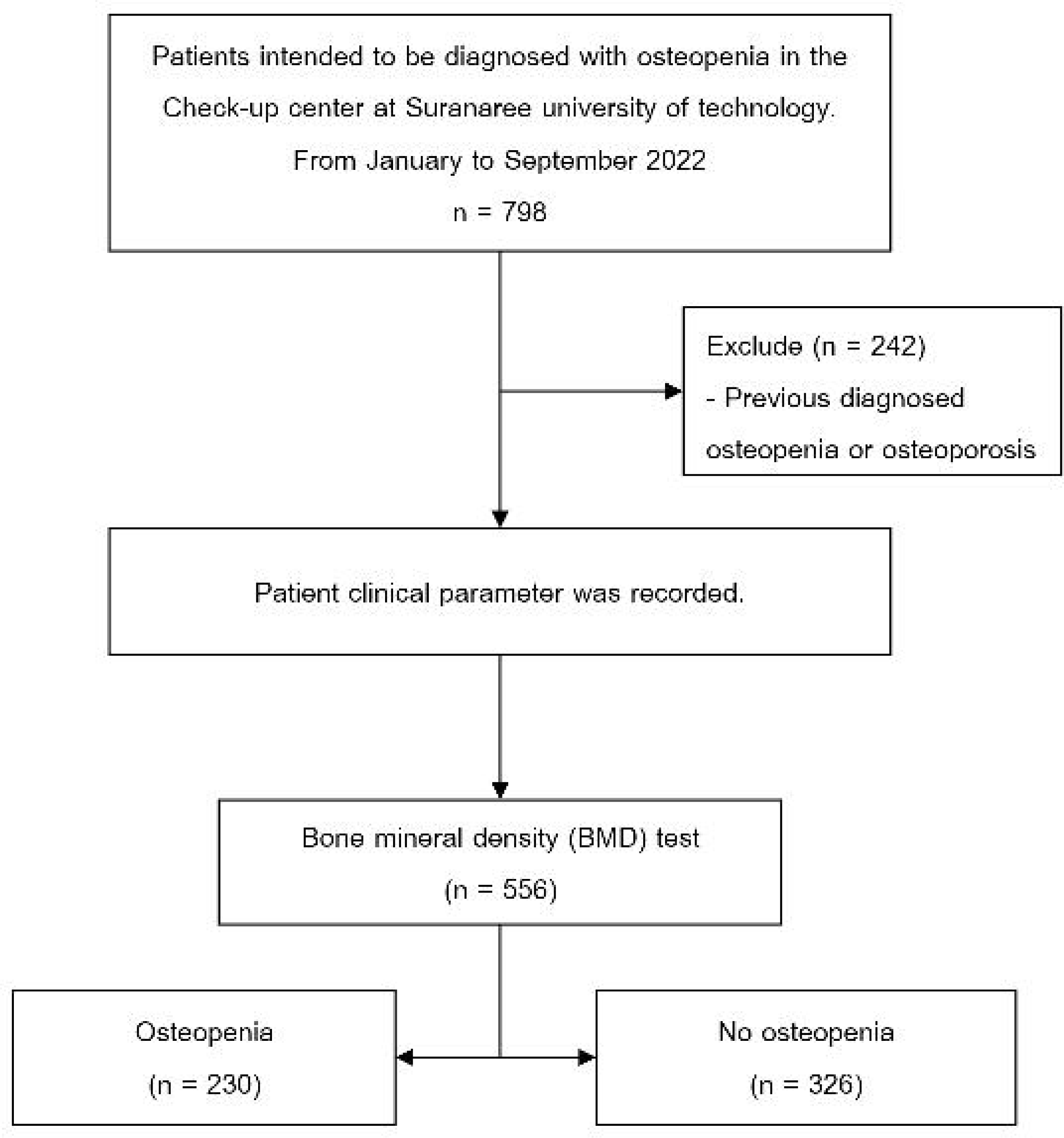
Study flow.

**Figure 2:**
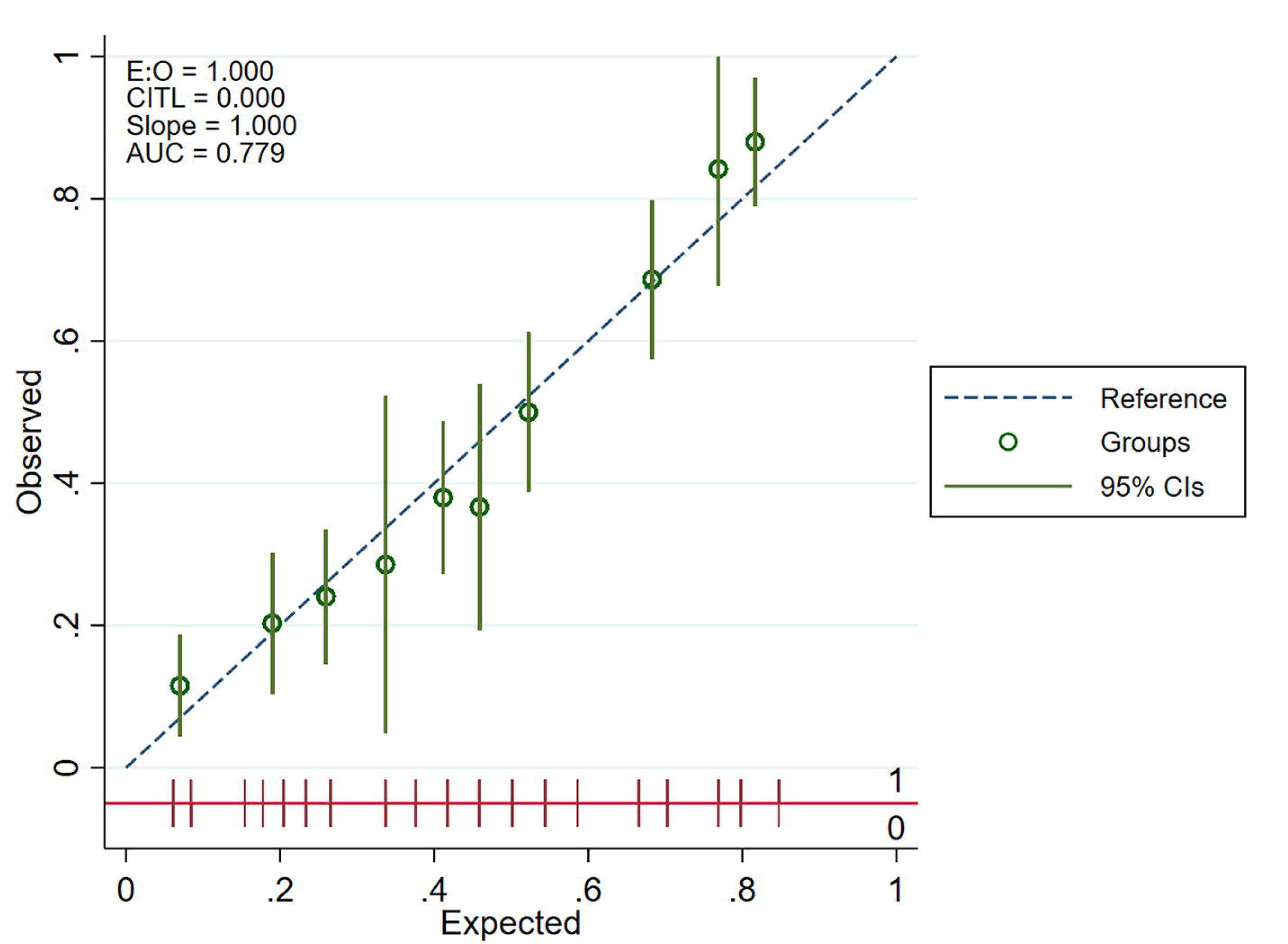
Calibration plot for model predicted risk for osteopenia versus actual risk.

**Figure 3:**
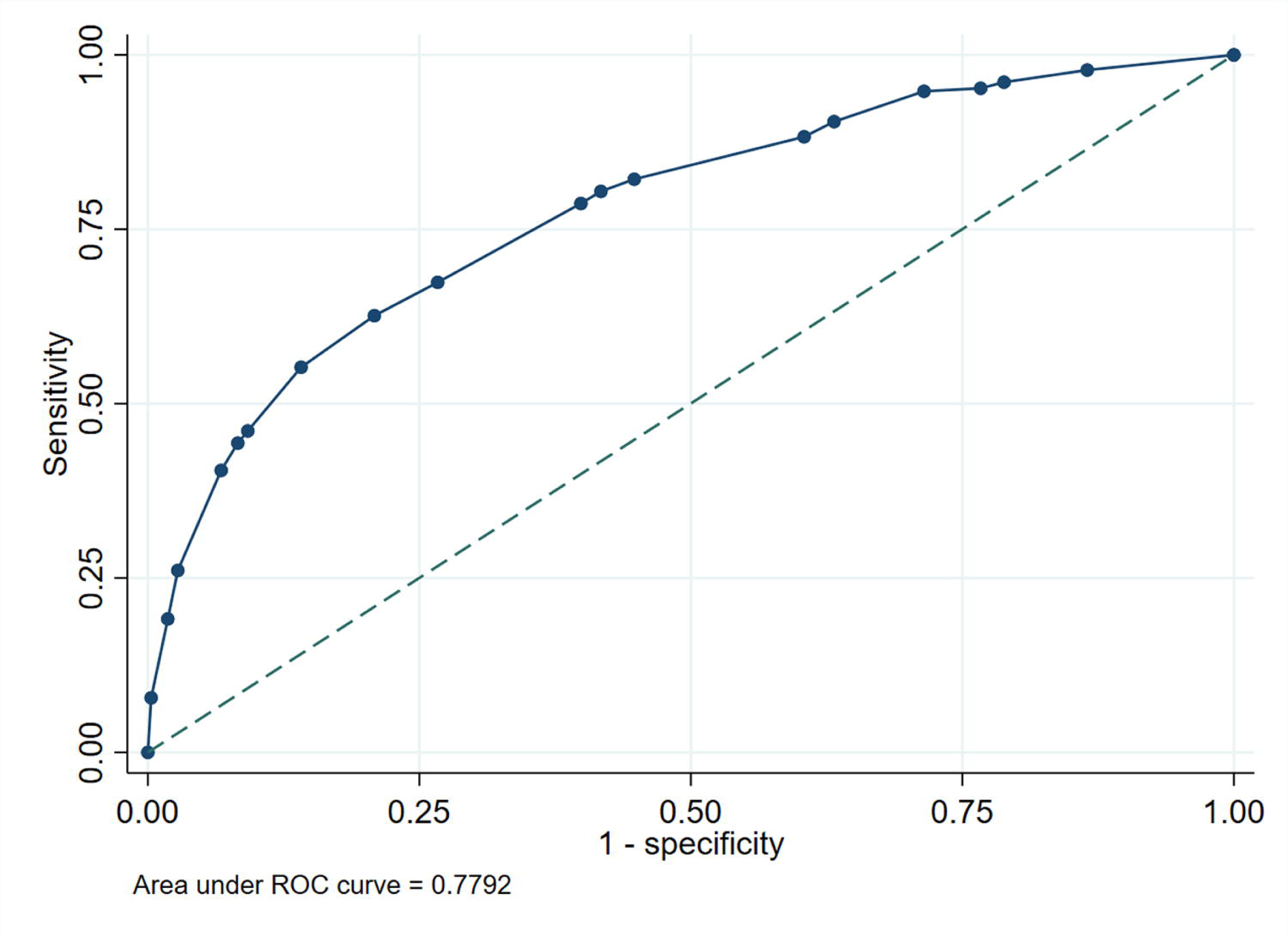
Discrimination performance of the newly developed model, using clinical characteristics to classify patients with normal and low bone mass density.

**Figure 4:**
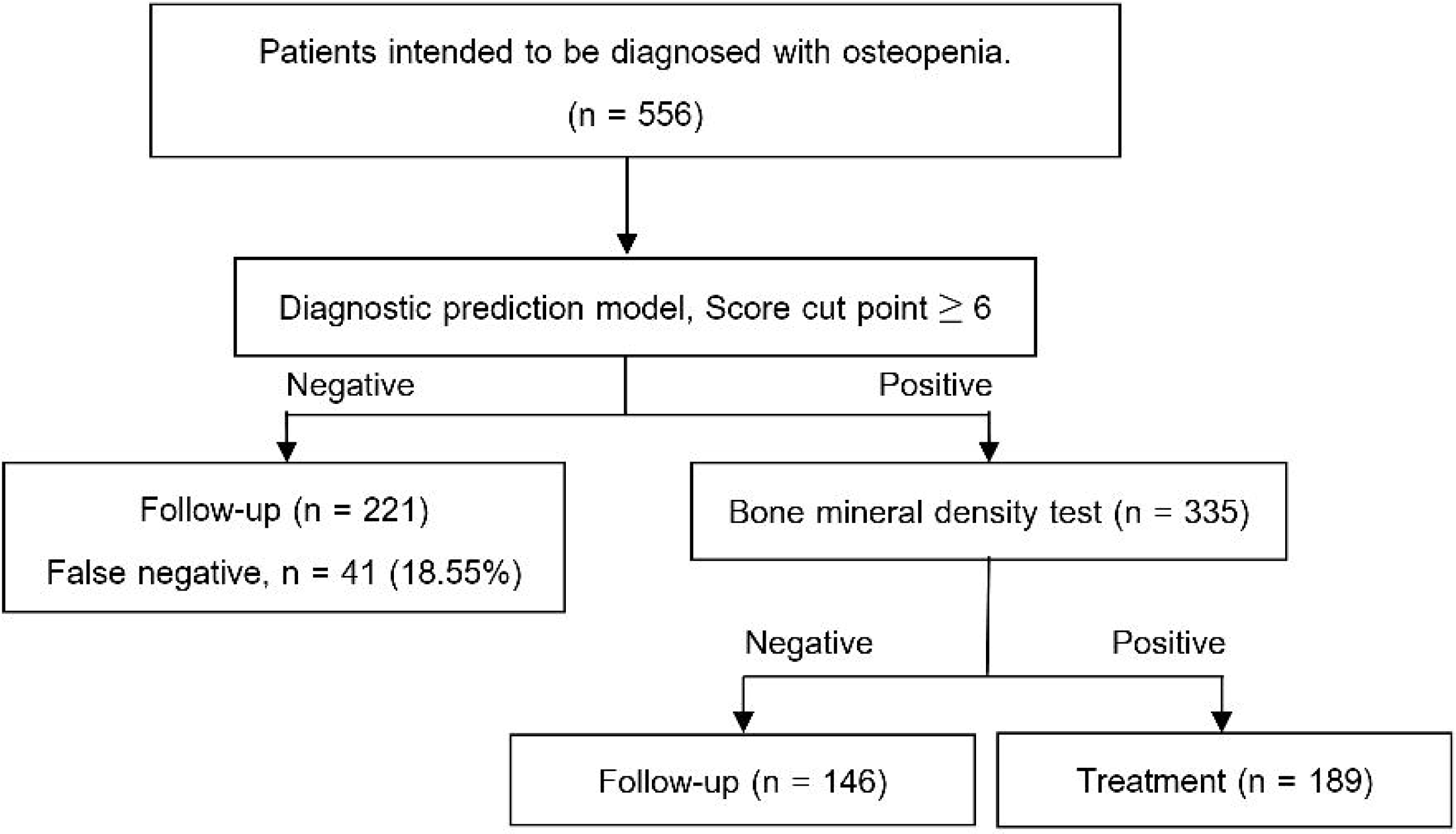
Clinical utility.

The process of internal validation was conducted by utilizing a bootstrap resampling technique with 1000 repeats. Following the adjustment for optimism in discrimination, the bootstrap analysis produced an area under the receiver operating characteristic curve (AuROC) of 0.768 (95% CI 0.73-0.81), indicates a good ability and a significant level of agreement between the estimated and observed probabilities of risk (Supplementary material).

### Simple score cut point identification

A very high sensitivity of 94.8% (95% CI 91.1-97.3) was observed at a cut point of ≥4, while the specificity was 28.5% (95% CI 23.7-33.8). At a score of ≥10 % (95% CI 88.2-94.5), but the sensitivity was lower at 44.3% (95% CI 37.8-51.0). The best cut point was determined by achieving a balance between sensitivity and specificity. For a score of 6 or above, the sensitivity was 82.2% (95% CI 76.6-86.9), and the specificity was 55.2% (95% CI 49.6-60.7), with a likelihood ratio of positive result of 1.83 (95% CI 1.60-2.10). The positive predictive value was 56.4% (95% CI 50.9-61.8), whereas the negative predictive value was 81.4% (95% CI 75.7-86.3) (Table 4).

**Table 4:** Selection of score cut-off point with sensitivity, specificity, LHR+, PPV, NPV, and along with 95% confidence interval.

| Score cut point | Sensitivity (%) | Specificity (%) | LHR+ (%) | PPV (%) | NPV (%) |
| --- | --- | --- | --- | --- | --- |
| ≥ 4 | 94.8 (91.1-97.3) | 28.5 (23.7-33.8) | 1.33 (1.23-1.43) | 48.3 (43.6-53.1) | 88.6 (80.9-94.0) |
| ≥ 6 | 82.2 (76.6-86.9) | 55.2 (49.6-60.7) | 1.83 (1.60-2.10) | 56.4 (50.9-61.8) | 81.4 (75.7-86.3) |
| ≥ 10 | 44.3 (37.8-51.0) | 91.7 (88.2-94.5) | 5.35 (3.63-7.90) | 79.1 (71.0-85.7) | 70.0 (65.4-74.3) |

Abbreviation: LHR+, likelihood ratio for positive test; PPV, positive predictive value; NPV, negative predictive value

### Clinical utility

In the context of triaging patients for osteopenia, the newly developed diagnostic prediction rule was applied. 32.37% (180 patients out of 556) were true negatives, this model can reduce the unnecessary BMD tests. Among the 211 patients who scored negatively (simple score below 6), 41 patients (18.55%) were false negatives for osteopenia, and only 2 patients (0.36% of all patients) were diagnosed with osteoporosis. On the other hand, among those with a positive score test (simple score above 6), 146 patients had normal BMD and were recommended for follow-up, while 189 patients were confirmed to have osteopenia and received medical treatment according to the treatment guidelines.

## Discussion

Despite the limitations, the current study managed to establish a diagnostic rule for predicting patients with osteopenia, which could strengthen an early diagnosis and the treatment of patients in osteopenia. This finding is highly relevant because our hospital faces a significant burden of bone density testing and a high prevalence of osteopenia. Moreover, the application of a predictive diagnosis logistic regression model, as conducted in the current study, also supports the efforts of other studies to implement more predictive analytics in clinical settings[20].

Our results emphasize the importance of a multi-factor approach in novel predictive model formulation. Specifically, our model uses the same age, sex, and weight as the known risk factors, established to predict osteopenia and osteoporosis, proven to be significant in other studies.[21] Similarly, the use of backward elimination for the purpose of precise identification of the factors allowed reducing the insignificant height and BMI from the model, as these variables have negligible impact on the model’s predictability of outcomes in our patients.[22]

The calibration and discrimination results of our model are satisfactory and being confirmed. The results of the Hosmer-Lemeshow test and AuROC testify to the model’s effective prediction of true osteopenia cases. This is confirmed by the literature on the appropriateness of calculating these metrics to check how well a diagnostic tool performs in clinical epidemiology and diagnostics.[23]

Furthermore, the use of this model demonstrated its significant clinical use. The prioritization of resources based on a simple score, which identifies high and low-risk patients, may also lower the risk of untreated osteopenia transitioning to osteoporosis and adhere to WHO, who suggests that more tests should be conducted on populations at higher risks.[24, 25]

Although the FRAX score is currently used in predicting the 10-year probability of hip fracture[26, 27], a study by Teeratakulpisarn et al[28]. reports that even though there is concordance between the 10-year probability of hip fractures for FRAX scores with and without BMD, this concordance declines in elderly and osteoporotic participants, and in those with FRAX scores without BMD. Therefore, to achieve higher accuracy, it is advisable to undergo BMD testing.

In 2001, Koh LK et al.[29] designed a simple tool to categorize postmenopausal Asian women (OSTA score). They utilized a questionnaire to identify those in the cohort with osteoporosis, defined as BMD T-scores ≤ −2.5 and use multivariable logistic regression analysis. The tool had a good performance with an area under the ROC curve of 0.79. Subsequently, it showed sensitivity of 91%, specificity of 45%, among others.[29] Additionally, upon a validation in Thai population, OSTA score presented sensitivity of 51.7% and specificity of 77.4% with a false negative rate of ∼20% The OSTA risk classification system showed that high and medium-risk patients were significantly more likely to sustain injuries in falls and have different femoral bone fractures patterns compared to low-risk patients. Machine learning models particularly artificial neural networks offer another opportunity to predict low BMD. Comparison of both ANN models to logistic regression models to predict low BMD had no significantly different in performance for either the femoral neck or lumbar spine.[34] Although the OSTA score performs well within the Thai population and particularly among postmenopausal women, however, it can be limited use to the general population.

Our model’s stability and reliability were internal validation via bootstrapping, process accounting for the potential optimism that can compromise prediction models developed in narrow or specific populations. All of these methods make our research more reliable and can be used to apply to similar settings with limited health care resources.

The study also has its limitations. In particular, the specificity of the model at some cut points was insufficient for real-life applications, resulting in overdiagnosis and overtreatment. This compromise between sensitivity and specificity is common in the development of diagnostic instruments, and it needs to be adjusted depending on the costs and risks of the disease. For future research, it is possible not only to include new, more prognostic factors but also to progress statistical instruments, such as machine learning. Further external validation is also required before adopting this model in other settings.

## Conclusion

The development of a diagnostic prediction rule for osteopenia in a resource-limited context is a major progress in the field of bone health management. This instrument is likely to enhance patient prognosis and maximize the use of available healthcare resources by detecting and offering timely therapeutic treatment to those at risk.

## Data Availability

All relevant data are within the manuscript and its Supporting Information files.

## Acknowledgement

This paper was supported by Suranaree University of Technology.

## Funding

None.

## Conflict of interest

None.

## Author contribution statement

NK and TJ contributed to all parts of the research.PT, KP, and PT focused on discussing, reviewing, and editing the manuscript.

## Notes

### Competing Interest Statement

The authors have declared no competing interest.

### Funding Statement

The author(s) received no specific funding for this work.

### Author Declarations

The study protocol received approval from the Institutional Review Board of Suranaree University of Technology, with approval number COA No.32/2567.

### Summary of Updates

Section on ethical consideration updated to clarify the date that accessed to patients data.

