## Supplementary material for "Development and Validation of a Diagnostic Prediction Rule for Osteopenia"

| **Apparent performance** | | | | |
| --- | --- | --- | --- | --- |
|  |  | 95% Confidence interval | | |
| Overall: |  |  |  |  |
| Brier scaled (%) | 23.7 |  |  |  |
| Discrimination: |  |  |  |  |
| C-Statistic | 0.780 | 0.741 | - | 0.819 |
| Calibration: |  |  |  |  |
| E:O ratio | 1.000 |  |  |  |
| CITL | 0.000 | -0.192 | - | 0.192 |
| Slope | 1.000 | 0.803 | - | 1.197 |
| **Bootstrap performance (Optimism adjusted)** | | | | |
| Number of replications: 1000 | |  |  |  |
| Overall: |  |  |  |  |
| Brier scaled (%) | 21.1 |  |  |  |
| Discrimination: |  |  |  |  |
| C-Statistic | 0.768 | 0.731 | - | 0.806 |
| Calibration: |  |  |  |  |
| E:O ratio | 1.004 | 0.918 | - | 1.089 |
| CITL | -0.004 | -0.199 | - | 0.197 |
| Slope | 0.936 | 0.759 | - | 1.151 |

**Shrinkage factors**

| Heuristic Shrinkage | 0.935 |
| --- | --- |
| Bootstrap shrinkage | 0.936 |

**Number of times each variable is selected**

|  | Freq | % |
| --- | --- | --- |
| female: | 1000 | 100.0% |
| 1b.score_age: | 0 | 0.0% |
| 2.score_age: | 1000 | 100.0% |
| 3.score_age: | 1000 | 100.0% |
| 1b.score_weight2: | 0 | 0.0% |
| 2.score_weight2: | 409 | 40.9% |
| 3.score_weight2: | 409 | 40.9% |
| 4.score_weight2: | 409 | 40.9% |
| score_height: | 177 | 17.7% |
| score_bmi: | 663 | 66.3% |
| diabetes: | 911 | 91.1% |
| score_egfr: | 131 | 13.1% |
